# Ketamine-Assisted Psychotherapy for Generalized Anxiety Disorder: A Comprehensive Case Report with Integrated Neurophysiological Imaging Using Magnetoencephalography

**DOI:** 10.1101/2025.02.25.25322463

**Authors:** J Cohen, AC Reichelt, R Zamyadi, G Roberts, S Hardy, B Fernandes, SG Rhind, EC Lewis, V Bhat, M Palner, R Jetly, BT Dunkley

**Affiliations:** Pharmacology & Toxicology, University of Toronto, Toronto, Canada; Neurosciences & Mental Health, The Hospital for Sick Children Research Institute, Toronto, Canada; Western University, London, ON, Canada; University of Adelaide, Adelaide, SA, Australia; MYndspan Ltd, London, UK; Field Trip Health, Toronto, Canada; Defence Research & Development Canada – Toronto Research Centre, Toronto, Canada; Hospital for Sick Children, Department of Pediatrics, University of Toronto, Toronto, ON, Canada; North Toronto Neurology, Toronto, ON, Canada; Interventional Psychiatry Program, St Michael’s Hospital, Toronto, Canada; University of Ottawa, Ottawa, Canada; Institute of Medical Science, University of Toronto, Toronto, Canada; Department of Psychology, University of Toronto, Toronto, Canada; Diagnostic and Interventional Radiology, Hospital for Sick Children, Toronto, Canada; Department of Medical Imaging, University of Toronto, Toronto, Canada; School of Psychology, University of Nottingham, Nottingham, UK; Clinical Physiology and Nuclear Medicine, Dept. Of Clinical Research, University of Southern Denmark, Odense, Denmark; Department of Nuclear Medicine, Odense University Hospital, Odense, Denmark; Department of Psychiatry, University of Toronto, Toronto, Canada

**Author notes:** **corresponding author:**, Benjamin T. Dunkley PhD, Department of Diagnostic and Interventional Radiology, 555 University Ave, Toronto, Canada, M5G 1X8, ex 328817.

**Keywords:** ketamine, psychedelic-assisted psychotherapy, Magnetoencephalography, cognition, functional connectivity, neural oscillations

## Abstract

This detailed case report explores the application of ketamine-assisted psychotherapy (KAP) in the treatment of a male patient in their late 30’s with Generalized Anxiety Disorder (GAD) and depressive symptoms. The N-methyl-D-aspartate (NMDA) receptor antagonist ketamine represents a significant breakthrough in the treatment of mood and anxiety disorders due to its rapid and robust antidepressant effects. Preclinical studies demonstrate that ketamine promotes biological alterations in the brain, including enhancing neuroplasticity. However, no studies to date have examined the longitudinal effects of KAP using magnetoencephalography (MEG), a powerful functional neuroimaging modality. Resting state MEG (rsMEG) scanning allowed the exploration of the neural correlates of KAP-related changes in mood and anxiety symptoms, including the functional connectivity between brain networks involved in cognition and emotion regulation. In this case study, an adult male participant with moderate-severe GAD underwent two rsMEG scans and cognitive testing at baseline and after 4 of 6 sessions of a standard ketamine administration and 2 integration sessions, part of a protocol consisting of a total of six KAP sessions and four sessions of integration. We measured functional connectivity in 5 functional networks – default mode, attention, central executive, motor, and visual, as well as neural oscillatory activity. We saw functional connectivity increases in 4 of the 5 networks. This coincided with significant increases in cortical beta activity, a marker of inhibition, decrease in theta oscillations, reductions in GAD7 and PHQ9 scores, and improved attention. In summary, these findings emphasize the ability of rsMEG to detect KAP-induced brain network changes, offering a promising tool for identifying clinically relevant neural correlates that can both predict and monitor therapeutic outcomes via electrophysiological changes.

## Introduction

Anxiety disorders, including Generalized Anxiety Disorder (GAD), are common psychiatric conditions affecting about 12.7% of people each year in the US (Olfson et al., 2019). GAD can be characterized by incessant, and uncontrolled worrying. These disorders are often chronic, with many patients experiencing symptoms for years before seeking treatment. This delay increases healthcare utilization and reduces quality of life and general functioning (Bandelow and Michaelis, 2015). Anxiety disorders are estimated to cost more than $4 billion in workplace productivity in the US alone and often contribute to the development or worsening of other medical conditions (“Mental Illness in the Workplace | Psychological Disability Management,” 2016; Sartorius et al., 2015).

Diagnosing an anxiety disorder involves criteria from the International Classification of Diseases, 11th Revision (ICD-11), or the Diagnostic and Statistical Manual of Mental Disorders, 5th Edition (DSM-5) (Association, 2013; “ICD-11,” 2022). The diagnosis is based on the severity, frequency, and persistence of symptoms, and must show significant psychological distress and/or impairment in daily functioning (Association, 2013).

While approved pharmacotherapies for anxiety disorders exist - including SSRIs and benzodiazepines, these drugs have limited efficacy, can produce adverse effects such as sedation and dependence, and require daily compliance (Kolovos et al., 2017). Although many patients with anxiety disorders experience symptom reduction with current treatments (Morilak and Frazer, 2004), up to 50% do not fully respond (Dunlop and Nemeroff, 2007; Gaynes et al., 2009). As such, anxiety disorders remain a significant challenge in psychiatric and psychological practices - particularly in treatment resistant cases (Penninx et al., 2021), necessitating innovative approaches.

Sub-anaesthetic ketamine without psychotherapy has emerged as a promising treatment for suicide-ideation, major depression and anxiety disorders due to its rapid therapeutic effects (Andrade, 2017; Swainson et al., 2021; Zanos and Gould, 2018; Tully et al, 2022; Witt et al., 20.22). Following the administration of a subanaesthetic dose of ketamine, patients with mood and anxiety disorders are reported to experience enhanced insight and cognitive flexibility, leading to a reduction in depressive thoughts and emotions. This pharmacological effect is utilized in ketamine-assisted psychotherapy (KAP) frameworks, taking advantage of a therapeutic window facilitated by ketamine-induced meta-plasticity (Krystal et al., 2023). In combination, ketamine and psychotherapy aims to reframe negative cognitive schemas and narratives to promote durable effects on mood (Walsh et al., 2022; Garel et al., 2023).

Ketamine is an N-methyl-D-aspartate (NMDA) receptor antagonist that interacts with cortical inhibitory interneurons, leading to an augmentation of downstream glutamatergic neurotransmission (Luscher, Feng et al, 2020). This surge of excitatory activity induces upregulation of the expression of synaptic plasticity-associated proteins (Haile et al., 2014) and increased cortical excitation (Cornwell et al., 2012), associated with the subsequent reduction in mood disorder symptoms (Zanos and Gould, 2018). Treatment with ketamine reverses glutamate synaptic signalling neuropathology through restoration of synaptic efficacy and synaptic density (Krystal et al., 2023).

In addition, ketamine inhibits hyperpolarization-activated cyclic nucleotide-gated nonselective cation 1 (HCN1) channels that are highly expressed in the hippocampus, cortex, and cerebellum. HCN1 contributes to the generation of rhythmic cortical oscillations and is a therapeutic target for affective disorders (Chen et al., 2009, Shah et al. 2012).

Preclinical research suggests that exposure to chronic stress disrupts extracellular glutamate levels via glial cell dysfunction. This often leads to overstimulation of extrasynaptic NMDA receptors, downregulates glutamate synaptic function, contributes to synaptic pruning, and produces depression and anxiety-like behaviour in animals (Hardingham et al. 2010). Ketamine’s synaptic meta-plastic effects play a role in sustaining antidepressant outcomes, helping to reverse the structural and functional synaptic changes seen in individuals with GAD (Grunebaum et al., 2018). Moreover, the effects of ketamine on brain-derived neurotrophic factor (BDNF) and mTOR signalling pathways are mechanistically proposed to foster synaptogenesis and maintained neural circuit integrity (Martinowich et al., 2007; Zanos and Gould, 2018; Haile et al, 2014).

Non-invasive functional brain imaging has proven valuable in understanding network-based biological mechanisms underlying psychedelic-assisted therapeutic mechanisms. For example, large-scale network changes induced by psilocybin and observed through functional MRI (fMRI) in the context of major depressive disorder (MDD) have been shown to modulate the default mode (DMN), central executive (CEN), and salience networks—a collection of networks that, individually and through their dynamic interaction, are involved in self-referential and autobiographical thinking. Taken together, these findings begin to show how psilocybin may disrupt the maladaptive thought patterns that characterize depression (Kuburi et al., 2022).

Ketamine-induced changes have also been observed to modulate activity in the DMN (Zacharias et al., 2020). While fMRI is a powerful tool for understanding functional brain changes, it is limited to imaging an indirect correlate of neuroplasticity in the form of haemodynamics, and the slow temporal resolution means it cannot detect rapid neural activity known as neural oscillations. Neural oscillations - or brain waves - are driven by temporal fluctuations in the excitability of neurons, and physiological states driven by neurochemical processes (Berger, 1929). Neural oscillations offer a unique macroscopic window in neural functioning, with different speeds of oscillations being directly related to various aspects of cellular physiology, as well as states of brain excitation & inhibition.

While traditional focus centres on glutamate neurotransmission, recent research also highlights alterations in gamma-aminobutyric acid (GABA)ergic transmission in brain regions associated with psychiatric pathophysiology as a potential contributor to ketamine’s antidepressant action (Silberbauer et al., 2020). These complexities underscore the need to bridge the translational gap between laboratory findings and clinical applications. Given the limitations of a single-case study, the contribution of this paper to the academic literature and the broader understanding of psychedelics/ketamine should be framed cautiously. However, the findings could potentially contribute to a more nuanced understanding of ketamine’s role in treating GAD and its potential implications for the future of psychiatric care (Sholevar et. Al, 2025). This case report aims to contribute to the medical literature by presenting a detailed analysis of both psychometric test scores, cognitive assessments, and neurophysiological outcomes measured by resting-state magnetoencephalography in a patient with GAD who underwent KAP.

## Methods

### Case Description & Diagnostic Assessment

#### Patient Information

The patient is a male in their late 30’s, with moderate-severe GAD, accompanied with depressive symptoms. He was diagnosed in 2021, through a thorough diagnostic evaluation which included psychiatric interviews, symptom rating scales, and an exploration of the patient’s treatment history by a psychiatrist specializing in mood and anxiety disorders.

Primary symptoms were irritability, insomnia and mental fog, with difficulty concentrating. The symptoms had been present for approximately 7 years before the formal diagnosis. Overall, the attending psychiatrist deemed the attitude of the patient as cooperative, affect was fully ranged and appropriate, with his thoughts being goal-directed and organized. His thought content was unremarkable with no suicidal ideas, intent, or plan. There was no evidence of any gross or severe cognitive or perceptual disturbance and insight was fair. At the time, the patient was deemed not to meet the criteria for resistant psychiatric illness and should not pursue ketamine treatment at that time. The patient was subsequently placed on SSRIs (Fluoxetine, 40mg/day) in June 2022 but was non-responsive and negative side effects (changes in weight and affect) resulted in a tapered withdrawal from pharmacotherapy in October 2022.

A second psychiatric assessment in November 2022 by a different psychiatrist deemed that the patient still met the criteria for moderate to severe GAD and that given the non-response to SSRIs and psychotherapy, was now eligible and a good candidate for KAP. The following contraindications were ruled out: Acute suicidality, significant Borderline Personality Disorder, ongoing substance abuse/use disorder, Manic/Hypomanic, and Psychosis. The following physical issues were also not present: severe sleep apnea, severe heart/lung disease, and active or past brain injury.

The patient also had a remote clinical history of transient Acute Stress Disorder from an adolescent traumatic episode but had otherwise recovered within 3 months of that incident with no specific intervention. The patient has no history of acquired brain injury, including mild traumatic brain injury/concussion, nor any history of participating in significant amounts of contact sports associated with repetitive subconcussive head impacts.

#### Clinical Findings

The patient exhibited pervasive low mood, overthinking, and worry, especially related to health anxiety and catastrophizing, anhedonia, sleep disturbance/insomnia, irritability, and self-reported cognitive impairment, including subjective reports of poor working memory and attentional abilities. The patient’s cognitive symptom profile has significantly interfered with work / occupational demands and some recreational interests, with poor memory and attention reducing optimal work productivity and performance generally. Otherwise, the patient maintained necessary work duties, relationships, exercise regimes and self-care largely without a problem. Past interventions, including various pharmacotherapies including SSRIs (Fluoxetine) and psychotherapies, including periodic visits to a clinical psychologist and counsellor over the 7 years prior for a formal diagnosis, yielded limited success. Before commencing KAP, the patient had completely withdrawn from SSRIs approximately 3 months prior to the baseline session, which were used for a total of 5 months (June-October 2022).

### Imaging acquisition and analysis

Neuromagnetic field data were acquired using a CTF-151 MEG scanner (CTF MEG Neuro Innovations Inc. Coquitlam, BC, Canada), with the patient in the supine position. Synthetic third-order gradiometry using the scanner reference array was used to attenuate environmental magnetic interference. Five minutes of eyes open resting state data were recorded at a sampling frequency of 600 Hz, which was filtered using a 1-150 Hz bandpass FIR filter to remove low-frequency drifts and very high-frequency oscillations; additionally, the data were notch filtered at 60 Hz to remove power line interference. These data were then downsampled to 300 Hz for further processing. Measurements of head location were acquired continuously during scanning using head position indicators placed at the nasion, and left and right preauricular points. Over the entire run, the overall displacement was judged not to exceed 5mm from the patient’s starting position.

Filtered data were segmented into 10.24 second epochs, and any epochs with a peak-to-peak amplitude of 6000 fT or exhibiting myogenic high-frequency activity were also excluded. 25 epochs were ultimately included for further analysis; this number was the same for both pre- and post-scans. Eye blinks and cardiac components were extracted from the data using independent component analysis and were automatically labelled for removal using a proprietary classification algorithm.

Data were subsequently source-localized using an LCMV beamformer. This technique combines a forward model of neural currents (generated using a template model of the inner skull) with the data covariance to adaptively estimate source currents. Source time courses (STC) were computed at a total of 122 locations: 78 locations using centroids from the AAL atlas (Gong et al., 2009) to estimate spectral power and 44 network nodes for functional connectivity (FC). For each STC, the data were scaled to their standard deviation.

Regional neural oscillatory power was measured across the brain across delta (1-3 Hz), theta (4-8 Hz), alpha (9-12 Hz), beta (13-29 Hz), and gamma (30-45 Hz) oscillations, which are implicated in inhibition and excitation and thalamocortical resonance. To infer regional activity, the power spectrum of the STC was estimated with the Welch method using 4096 samples per segment. Taking the mean power within each band of interest provided an estimate of relative magnitude for that band at a given location in the brain.

Functional connectivity was measured across 5 functional networks, each using a specific frequency to define functional connectivity – default mode (alpha 8-12Hz), attention (alpha 8-12Hz), central executive (alpha 8-12Hz), motor (beta 15-25 Hz), and visual (alpha 8-12Hz). Connectivity estimates were performed using the corrected amplitude envelope correlation method. Scanning was performed twice, the first was recorded prior to ketamine dosing (the baseline scan), and the second after the 4th dosing schedule was completed (follow-up scan), when symptoms had decreased to stable levels.

Oscillatory activity and connectivity analyses were compared against a normative dataset of 170 neurotypical controls (107 females, and 63 males). All observed FC and power values were mean-centred and scaled to the standard deviation of the normative group to provide a z-score for each observation in frequency and location: this effectively creates a regional map of z-scores for each scan. Subtracting the baseline z-values from the follow-up z-values generates a single map which represents neurophysiological changes between scans in units of population-level standard deviations.

### Therapeutic Intervention

The participant underwent a standardized CORE KAP protocol offered by Field Trip Health in January 2023. This protocol consisted of six ketamine dosing sessions, each lasting 90 minutes, over 3 weeks, with each session supervised and paired with psychotherapy. During each dosing session, the participant received sublingual racemic ketamine hydrochloride troches at doses detailed in **Table 1**. The dose was spat out after 10 minutes of swilling. Sublingual administration leads to the onset of acute ketamine effects within 10-15 minutes. The specific dosing regimen was used in Field Trip Health Clinics as of January 2023, which was prescribed by a MD of Psychiatry and treatment sessions were monitored by a registered nurse and licensed therapist in a clinical setting. During the administration sessions, the patient was supine in a chair with a weighted blanket, wore an eye mask, and listened to a variety of auditory accompaniments, including music, nature sounds, and synthesized auditory soundscapes.

**Table 1.** Dosing schedule for KAP.

| Schedule | Day | Dose (mg) |
| --- | --- | --- |
| Baseline MEG scan 1 | 0 | - |
| Ketamine 1 | 0 | 250 |
| Psychotherapy 1 | 2 | - |
| Ketamine 2 | 4 | 350 |
| Psychotherapy 2 | 6 | - |
| Ketamine 3 | 8 | 450 |
| Ketamine 4 | 11 | 550 |
| MEG scan 2 | 13 | - |
| Psychotherapy 3 | 14 | - |
| Ketamine 5 | 15 | 600 |
| Ketamine 6 | 18 | 700 |
| Psychotherapy 4 | 20 | - |

*This table outlines the dosing and psychotherapy sessions administered during the Ketamine-Assisted Psychotherapy (KAP) treatment regimen, including the specific ketamine doses and dates of administration. Psychotherapy sessions were scheduled in between ketamine administrations as indicated by the blank dose fields.*

**Table 2.** Studies showing neural effects of ketamine.

| Study | Year | Disorder | Ketamine Protocol | Key Finding | Neural Correlate |
| --- | --- | --- | --- | --- | --- |
| Abdallah et al. | 2017 | MDD | Single dose IV ketamine | Increased global brain connectivity in lateral PFC, caudate, and insula; normalization of dysconnectivity between PFC/subcortex and rest of brain | Potential mechanism for ketamine's antidepressant effects |
| Nugent et al. | 2020 | MDD | Pre- and post-KAP MEG scans | Reduced aberrant connectivity in DMN and CEN; increased coherence in SN | Large-scale brain networks involved in emotion regulation and attentional processing |
| Tian et al. | 2023 | N/A | Intracranial recordings | Increased gamma oscillations in prefrontal cortex and hippocampus | Implicated in ketamine's antidepressant effects |
| Rengasamy et al. | 2024 | Treatment-resistant depression | Multiple-dose IV ketamine | Greater prefrontal/limbic connectivity post-ketamine treatment; lower baseline connectivity predicted better response | Importance of baseline connectivity in predicting treatment response |
| Current Study | 2025 | GAD | Oral | Similar findings to previous studies, suggesting common neural substrates underlying various psychiatric disorders | Reinforces the role of ketamine in modulating large-scale brain networks and its potential for treating diverse mental health conditions |

Therapeutic support immediately before and after the ketamine dosing session was offered by the attending psychotherapist to help set intentions, facilitate emotional processing, and evaluate the session. Additionally, four separate 60-minute psychotherapy-based follow-up integration sessions held on different days to the ketamine sessions were also conducted via one-to-one telehealth with a licensed therapist over the course of 3 weeks. Integrative psychotherapeutic approaches were manualized, including structured guidelines for clinicians to follow to ensure standardized and effective treatment. However, since the treatment was client-centered, there was a degree of flexibility to accommodate the diverse needs, backgrounds, and presentations of the clients who accessed the clinic. This balance between structure and adaptability allowed clinicians to maintain consistency while tailoring the approach to each individual’s unique circumstances. The integration sessions combined cognitive behavioural therapy, internal family systems, and mindfulness amongst others (Zarbo et al., 2016).

The dosing schedule was started at 250 mg, as per Field Trip Health Standard of Care for this protocol, and gradually increased based on patient feedback. The patient had prior experience with recreational psychedelics and was comfortable with the increasing dose over sessions.

### Cognitive assessments

Cognitive-behavioural assessments were administered using a tablet which included a battery of validated tasks (Creyos Ltd, https://creyos.com/online-cognitive-tasks), measuring sustained attention/concentration, logical & grammatical reasoning, short-term working memory, visuo-spatial attention, and visuo-spatial perceptual abilities. Performance was assessed against a normative database of more than 85,000 participants with the patient matched to the appropriate age range and sex. See **Supplementary Materials** for details.

### Psychometric assessments

The patient also completed self-report screener forms including the Patient Health Questionnaire 9 (PHQ-9) item scale (Kroenke et al, 2001; Kroenke et al, 2009) and the Generalised Anxiety Disorder 7 (GAD-7) item questionnaire (Spitzer et al, 2006) The PHQ9 uses nine questions to assess Major Depressive Disorder and it has been shown to have good internal consistency and validity. Higher scores for the PHQ-9 represent increased likelihood of Major Depressive Disorder. The GAD7 uses seven questions to assess for Generalized Anxiety Disorder and has been shown to have good psychometric properties. Higher scores for the GAD-7 indicates likelihood to meet the criteria for Generalized Anxiety Disorder. These scales were administered at approximately two weeks, one month and three months post-intervention.

## Results

### Psychometric assessments and clinical notes

As shown in **Figure 1**, KAP resulted in clinically meaningful and sustained reductions in severity of anxiety and depression symptoms. Improvements were noted rapidly and plateaued after the 3rd KAP session. These improvements were maintained at the 80-day follow-up, indicating a sustained therapeutic benefit from ketamine assisted therapy and psychotherapeutic integration sessions.

**Figure 1.**
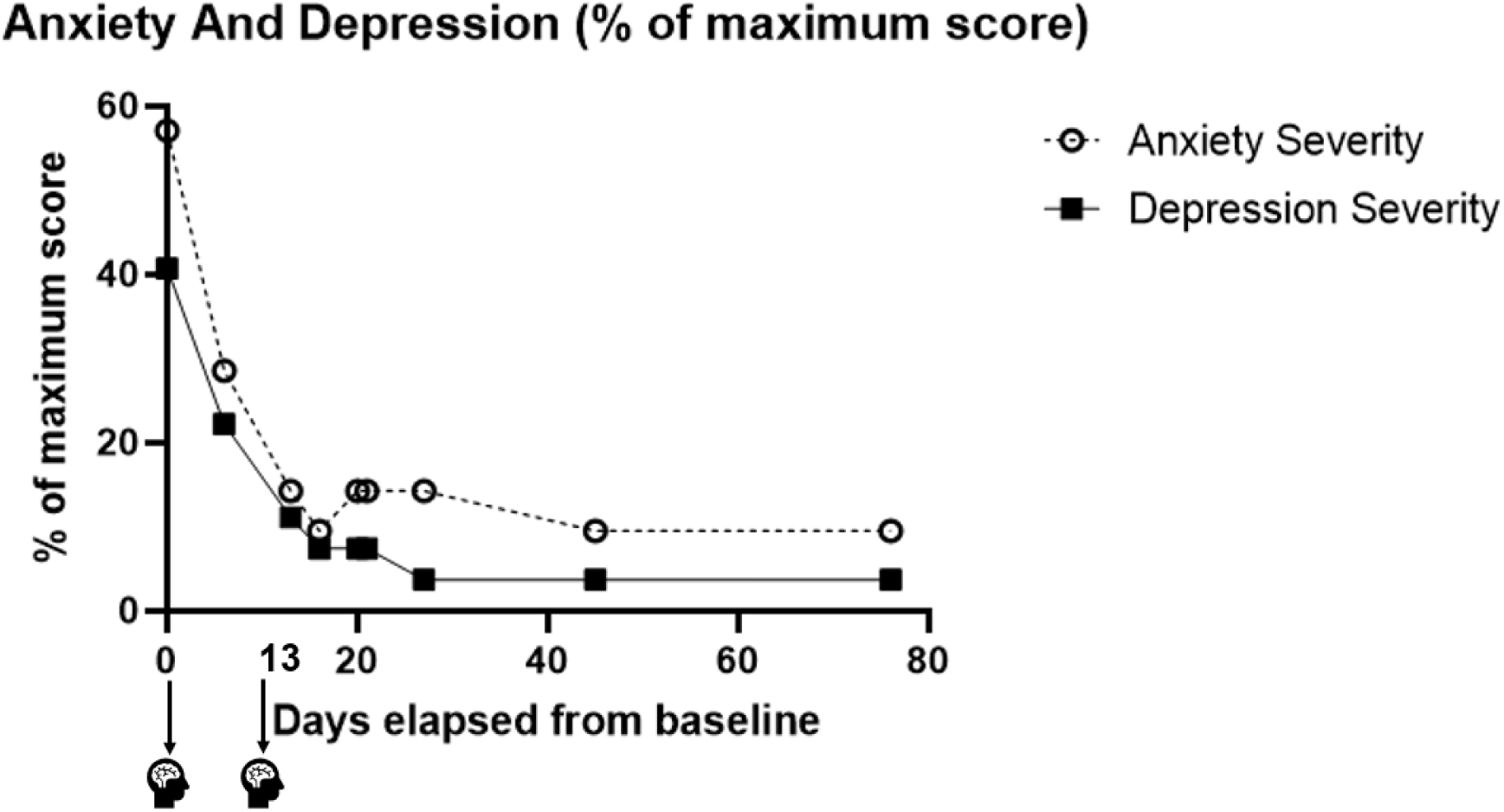
Self-reported changes in anxiety and depression symptoms over a three-month period. The x-axis represents time in days (Day 0 = Baseline, pre-KAP). The y-axis represents scores on the PHQ-9 (Patient Health Questionnaire-9) and GAD-7 (Generalized Anxiety Disorder-7) scales, expressed as a percentage of the maximum possible score (higher scores indicate greater symptom severity). Baseline MEG was recorded at on the same day and before beginning KAP therapy (Day 0, **Table 1**) and a scan was recorded after the 4^th^ ketamine and 2^nd^ integration session therapy (Day 13, **Table 1)**.

The final integration session included a reflection on the progress during treatments and the patient indicated that anxiety has decreased, in particular health anxiety, as well as feelings of hypervigilance and a change in perspective. The patient was reflective and understood the importance of mindfulness and being in the present moment. At the closure of therapy, the patient attested that they were able to be more introspective of their thoughts, behaviours and feelings, and reported increased engagement in meaningful activities.

### KAP was associated with changes in cortical oscillatory activity and network functional connectivity

#### Cortical oscillatory activity

rsMEG analysis showed changes in cortical oscillatory activity, with the largest changes occurring in the theta and beta band (**Figure 2**), for the most part, across the entire cortex. Theta oscillatory activity showed reductions between scan 1 (normalized z score=0.42 compared to normative database) to scan 2 (z score=-0.04, change in z score −0.46), while beta activity increased between scan 1 (z score=-0.27) and scan 2 (z score=0.23, change in z score +0.50).

**Figure 2.**
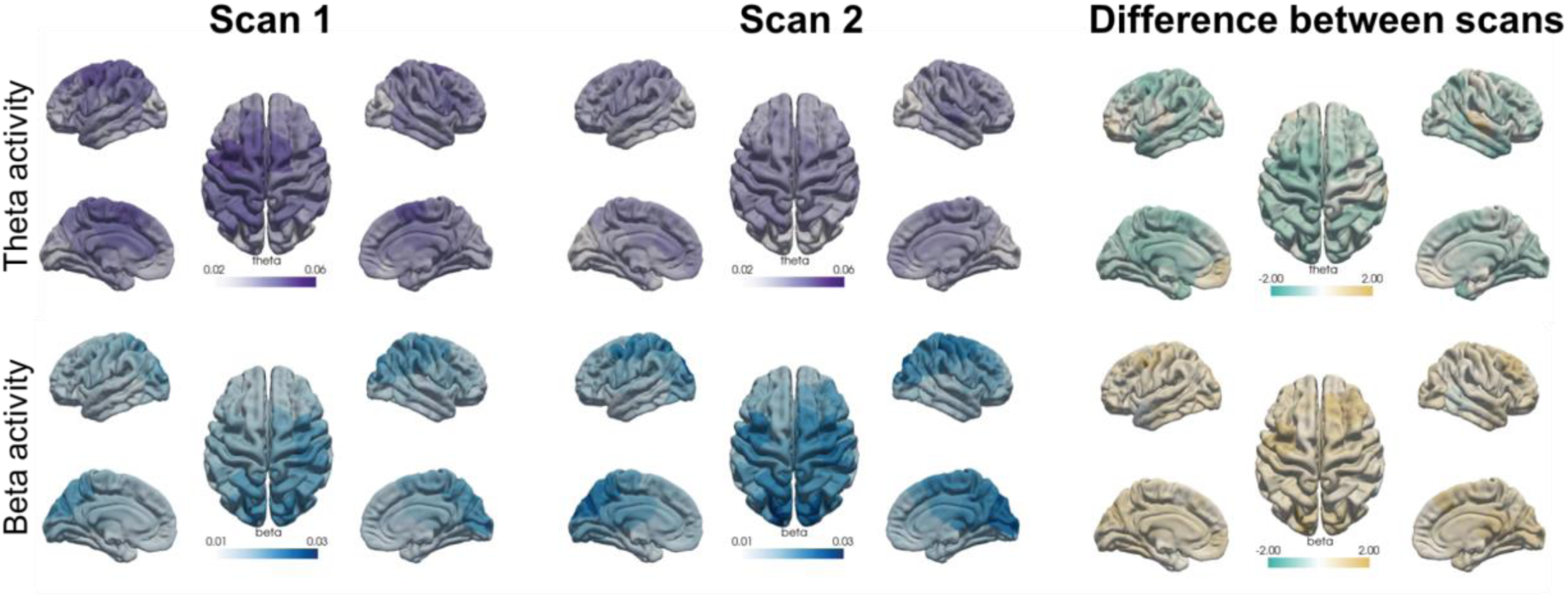
Regional neural activity changes were most pronounced in the theta and beta bands between scans, showing reductions in theta activity and increases in beta activity. Oscillatory analyses were compared against a normative dataset of 170 neurotypical controls (107 females and 63 males) with a mean age of ∼ 32 years.

#### Functional connectivity

rs-fcMEG analysis showed substantial changes in normalized functional connectivity in 4 of the 5 networks analysed (**Figure 4**). rs-fcMEG across all networks increased following KAP, including the CEN (from 10th percentile to 52nd percentile), visual (2nd to 12th percentile), DMN (15th to 57th percentile), attention network (15th to 55th percentile), motor network (18th to 64th percentile).

**Figure 3.**
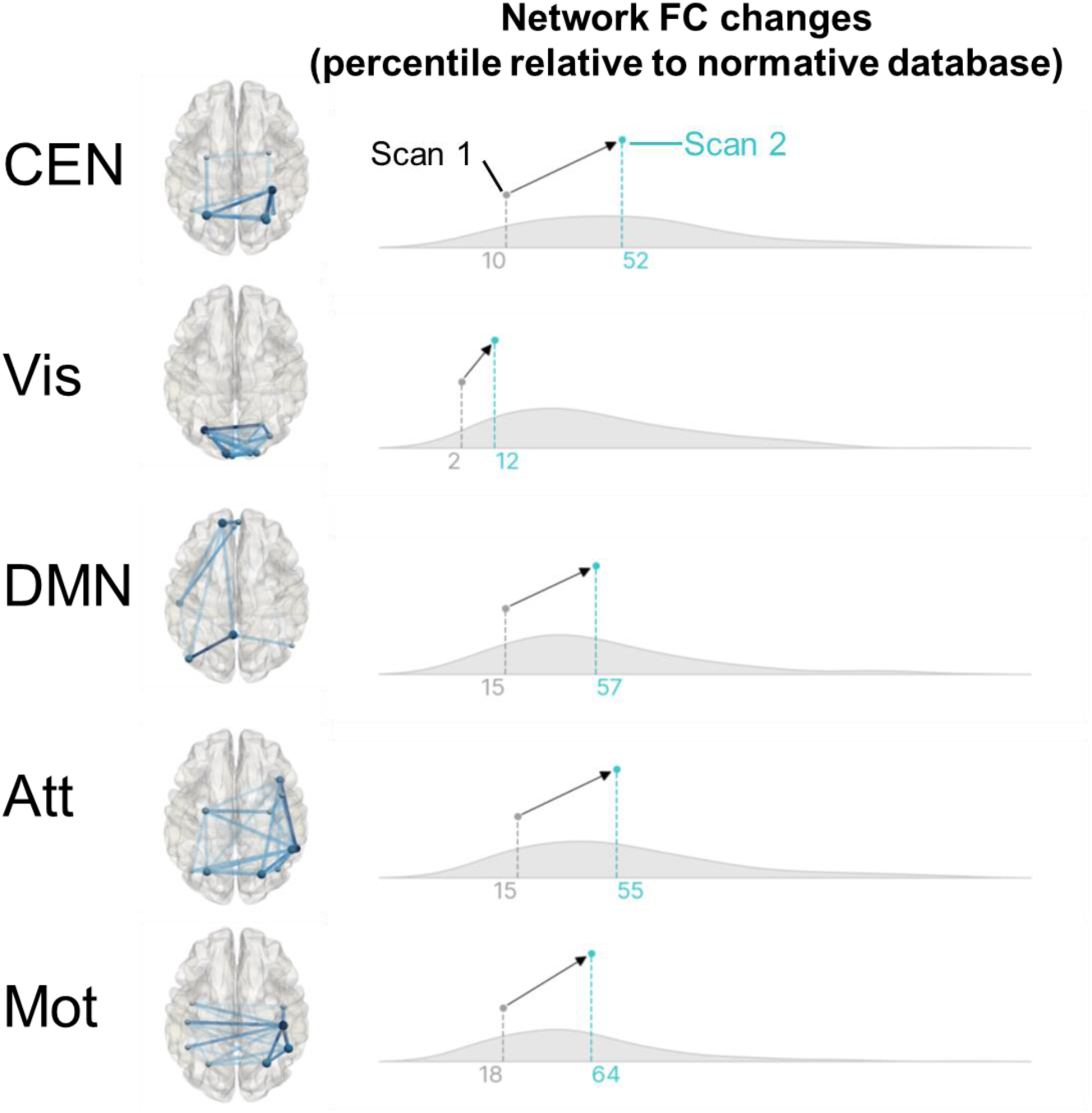
Within-network neural synchrony (or functional connectivity) was measured across 5 functional networks; default mode, attention, central executive, motor, and visual. Functional connectivity analyses were compared against a normative dataset of 170 neurotypical controls (107 females, and 63 males), with values indicating percentile (relative) placement to the neurotypical controls at scan 1 (grey) and scan 2 (blue values).

**Figure 4.**
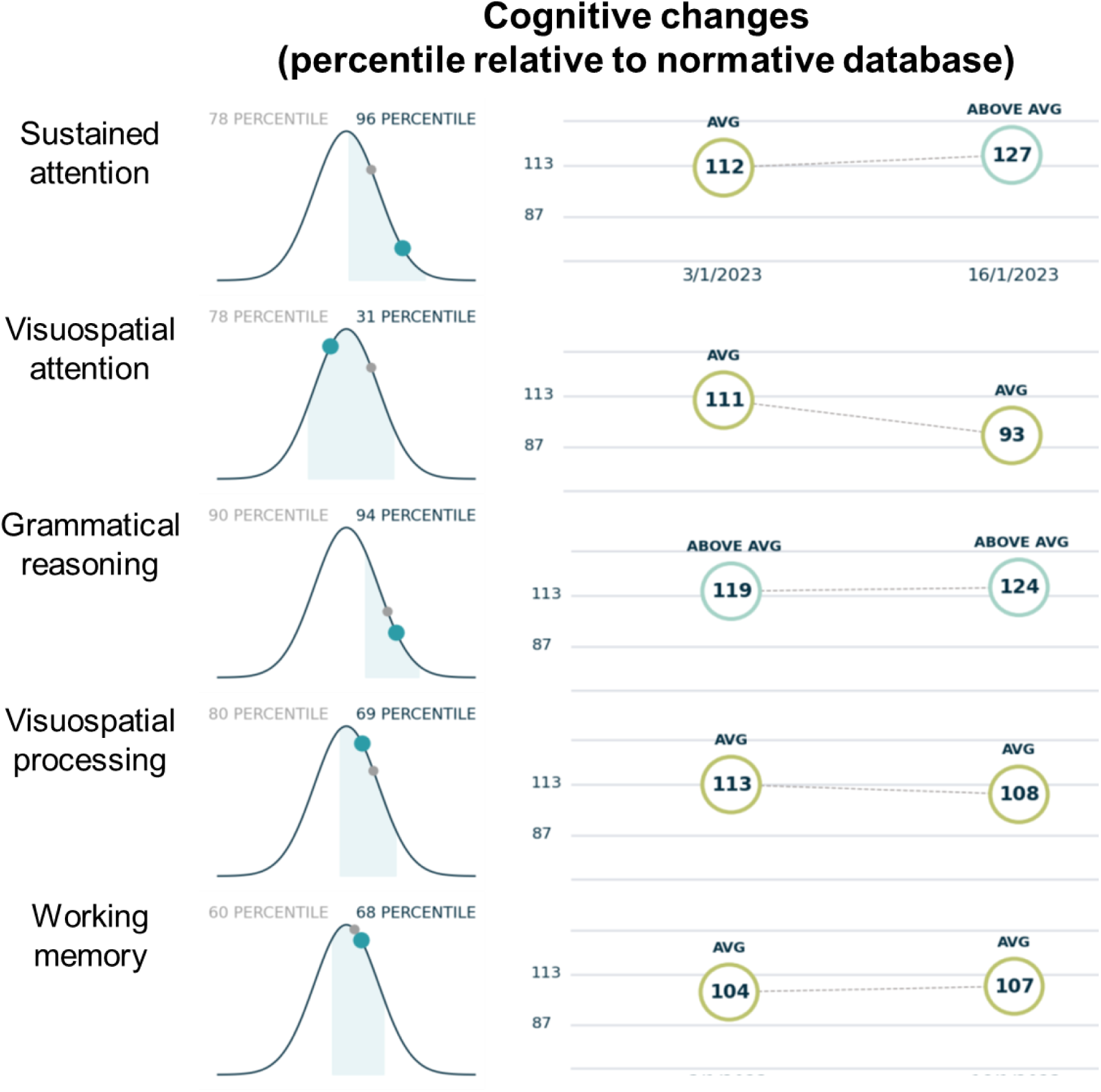
Longitudinal cognitive assessments across 5 domains, including sustained attention, visuospatial attention, logical/grammatical reasoning, visuospatial processing/discrimination, and working memory. Cognitive performance was compared against a normative database of more than 85,000 participants and presented as percentile (relative) placement to the database at scan 1 (grey) and scan 2 (blue values)

#### Cognitive assessments

Cognitive testing showed there were meaningful improvements in sustained attention between assessments (**Figure 4**), but decreases in visuospatial attention, whilst grammatical reasoning, visuospatial processing, and working memory remained relatively unchanged between assessments.

## Discussion

This case report provides a novel exploration of KAP for the treatment of GAD and depressive symptoms with neural correlates measured through rsMEG. The application of KAP represents an important step forward in psychiatric care, especially in patients unresponsive to traditional treatments (Sholevar et al, 2025). Previous research has demonstrated the rapid and robust antidepressant effects of ketamine (Krystal et al., 2013), largely attributed to its modulation of the NMDA receptor and the associated increase in metaplasticity (Carhart-Harris and Friston, 2019). This case study is the first to use rsMEG to map the oscillatory and functional connectivity changes that accompany symptom improvement in a patient with GAD, providing important insights into the potential neural mechanisms underlying the therapeutic effects of KAP.

The subject demonstrated changes in neural network connectivity associated with clinically meaningful reductions in depression and anxiety symptom scores and improvement in cognition with sustained duration. These findings were accompanied by significant changes in cortical oscillatory activity, particularly in the theta and beta frequency bands, observed through rsMEG. Theta oscillatory activity showed reductions (normalized z score decrease from 0.42 to 0.04), while beta activity exhibited increases (z score increase from −0.27 to 0.23), reflecting widespread neurophysiological changes across the cortex. The reduction in theta activity suggests a normalization of hyperactive emotional circuits, potentially dampening excessive limbic system activity (Curic et al., 2021). Beta activity increase may reflect connectivity strength, promoting cognitive flexibility and emotional regulation (Rivolta et al., 2015). Counter to expectations, the beta activity changes are opposite to the effects observed during *acute* ketamine infusion (Machado et al., 2022) which might reflect differences in mechanisms related to administration intervals. Curic et al. however, also observed increased beta activity post-administration which is related to positive increases in connectivity and cognitive flexibility (Curic et al., 2021; Rivolta et al., 2015). This perhaps represents a stable or adaptive neural state following intervention. In contrast, acute IV administration of ketamine induces a “glutamatergic surge” that disrupts glutamatergic signalling. This surge leads to transient cortical desynchronization, characterized by a reduction in beta activity alongside an enhancement of slower (theta/delta) and faster (gamma) oscillations (Machado et al., 2022).

These findings highlight the context-dependent nature of beta oscillations when attempting to understand their meaning clinically. The acute ketamine treatment period correlates with well-established patient reports of sensorimotor, emotional and cognitive perceptual changes likely permitted by the diminished stability and resultant dynamic reconfiguration of intra-network connectivity during exposure. When captured, this state appears to be reflected by a reduction in beta oscillatory activity as described by previous investigators (Machado et al., 2022). Our patient described the same subjective, clinical impact with respect to acute ketamine exposure and subsequently ongoing positive changes in mood and cognition associated with observable, significant increases in cortical beta activity and functional connectivity within 4 out of 5 large-scale networks measured after 4 ketamine sessions when clinical symptoms had reduced significantly. This opposing state of increased beta activity, captured in the period following exposure, may then be a marker of persisting, stable, intra-network reconfiguration, Thus, the tendency of beta oscillations to change in relation to exposure is an intriguing marker of treatment response to ketamine in the context of anxiety disorders. This also highlights the need to consider timing and intervention context when interpreting neural oscillations, further underscoring the importance of the role of beta oscillations in regulating cognitive and emotional processes.

While increased beta activity is associated with inhibition, stability, and enhanced functional coupling across brain networks, its reduction during ketamine infusion reflects a transient disruption in the excitation-inhibition balance, characterized by decreased inhibition and a relative increase in excitation. This shift is integral to ketamine’s psychotherapeutic mechanism. These dynamics highlight the importance of considering timing and intervention context when interpreting neural oscillations, highlighting the critical role of beta oscillations in regulating cognitive and emotional processes across different treatment modalities.

The observed oscillatory changes align with the hypothesized role of ketamine in promoting synaptic plasticity through NMDA receptor modulation (Cornwell et al., 2012; Krystal et al., 2013). Together, these results highlight the potential of rsMEG to advance our understanding of the neurophysiological underpinnings of KAP and its therapeutic mechanisms, while also providing a foundation for future research to refine treatment protocols and identify objective markers of clinical response.

### Patient & Therapist Perspective

> The patient’s perspective is integrated, highlighting his experiences, insights gained through the therapy, and reflections on the overall impact of KAP on his well-being. At referral, presents to our clinic today with a history of GAD for a number of years. The patient mentions that he grew up in a household with a very positive family upbringing. About 2016, the patient started to experience heightened states of stress, sleep disturbances, and irritability – his neuropsychiatric symptoms were triggered by stressors. The patient’s symptoms worsened to the point that they became debilitating - he couldn’t focus during the day.

> The patient was doing reasonably well having tried multiple approaches to help mitigate his neuropsychiatric symptoms, including life-style modifications, psychotherapy, neurofeedback, and pharmacotherapy. But these measures have not helped with continued GAD. The patient’s chief complaints were states of generalised worry, intrusive thoughts, and an inability to relax, poor sleep quality and quantity (associated with initiation and maintenance problems). His coping strategies at this time include exercising, implementing sleep hygiene strategies, and continuation of his talk therapy (including addiction counselling). However, the patient still felt that he has not been able to mitigate his daily states of intrusive thoughts and racing mind. He has not been able to fully enjoy life, unable to live in the present as he is constantly worried about the future.

> Prior pharmacotherapy with Prozac and Zopiclone, used for 5 months, was discontinued due to side effects he experienced – including exacerbation of GERD and weight gain. Intermittent psychotherapy for 8 years ago with numerous Clinical Psychologists, Counsellors, and a Psychotherapist managed anxiety, but did not address the root cause. Although the patient was unable to pinpoint the main source of his anxiety, he tends to overthink about work and life, thinks of hypothetical situations in his mind that he believes is unlikely to happen, and is unable to relax. The patient endorsed being restless, fatigued, and has excessive anxiety and worry, impaired concentration, irritability, and difficulty sleeping. The patient endorses that he has good support systems. He is interested in KAP as he has tried psilocybin approximately 8 times to date, last use was 2 years ago. Overall, he had positive experiences and noticed benefits with this. He has also looked into the data and research of KAP and believes this could be beneficial for him

> After his sessions, the patient reflected on his 6 KAT and 4 integration sessions, which he stated has revealed to him how his health anxiety had decreased substantially. Both the patient and the integration therapist reflected on the impact KAP and the role it had to play in reducing hyper-vigilance as well as the perspective change that has helped reduce the anxiety around health. The patient and therapist discussed the importance of accepting the natural decline of functioning that comes with ageing, but also being mindful of and living within the present moment. The patient and therapist discussed the narrative around finishing KAP sessions, with the patient stating that KAP provided an introduction to himself and that he was able to be more introspective of his thoughts, behaviours and feelings and overall more reflective.

Affective disorders are characterised by dysfunctions in attention and difficulties in concentration and decision making. It has been shown that patients with anxiety disorders show impairments in working memory and restrain irrelevant information processing, leading to cognitive and attentional deficits. This profile of neurocognitive impairment in anxiety and mood disorders may be attributable to a deficit in the efficacy of network level activity, with the CEN’s role in human working memory (Moon & Jeong, 2017) well established. In this case study, the subject reported impairments in attention and working memory, despite performing at a comparatively high level in all cognitive assessments. Improved sustained attention performance was observed post-KAP, which indicates that the subject experienced functional improvements in this cognitive domain. This observation is supported by previous studies that have shown low dose sublingual ketamine produced rapid and sustained effects in improving mood, cognition and sleep in over 75% patients in their study cohort with refractory unipolar or bipolar depression (Lara et al, 2013).

With rsMEG, we explored the objective neural correlates of psychedelic-related changes in mood and anxiety symptoms, including the functional connectivity within brain networks involved in cognition. In the subject with GAD, brain changes between baseline at post-treatment sessions suggests *increased inhibition* demonstrated by increases in beta-band frequency oscillations. rsMEG scanning shows promise as an objective biomarker in tracking neural correlates altered by psychedelics to help monitor therapeutic outcomes in patients.

Despite the promising findings presented in this case report, several limitations should be acknowledged. The single-case design limits the generalizability of findings to broader populations, emphasising the need for replication in larger, more diverse samples. Additionally, the lack of a control condition limits the ability to draw definitive conclusions regarding the specific efficacy of KAP compared to alternative interventions.

### Conclusion

This comprehensive case report provides a detailed examination of KAP in the treatment of GAD, adhering to CARE guidelines. In this case report, neural correlates via MEG imaging correspond with improved clinical outcomes. MEG can be a powerful and meaningful tool when monitoring neural networks and connectivity in tandem with a pharmacological or psychological treatment modality. This report contributes to the evolving literature on psychedelic-assisted therapies and underscores the need for further research to examine changes in neural networks and explore the broader applications of KAP in mainstream psychiatric practice.

Future research employing rigorous study designs, including randomised controlled trials with active comparator groups, is warranted to establish the relative efficacy and safety of KAP in GAD. The findings presented in this case report contribute to the growing body of evidence supporting the therapeutic utility of KAP in GAD. This report underscores the relevance of these findings within the field of personalised psychedelic therapy with implications for improving outcomes and quality of life for individuals affected by GAD and, emphasising the need for further research to elucidate the underlying mechanisms of action, optimise treatment protocols, and facilitate the integration of KAP into routine clinical practice.

## Data availability statement

The original contributions presented in the study are included in the article/supplementary material, further inquiries can be directed to the corresponding author.

## Ethics statement

Written informed consent was obtained from the individual for the publication of any data included in this article. The authors declare that information from this case study was collected in accordance with the standard of care.

## Author contributions

JC: Conceptualization, Data curation, Formal analysis, Investigation, Methodology, Visualization, Writing – original draft, Writing – review & editing.

ACR: Conceptualization, Data curation, Formal analysis, Investigation, Methodology, Visualization, Writing – original draft, Writing – review & editing.

GR: Data curation, Formal analysis, Methodology, Visualization, Writing – original draft, Writing – review & editing.

SH: Data curation, Formal analysis, Methodology, Visualization, Writing – original draft, Writing – review & editing.

BF: Writing – original draft, Writing – review & editing

SR: Writing – original draft, Writing – review & editing

VB: Writing – original draft, Writing – review & editing

RJ: Writing – original draft, Writing – review & editing

BTD: Conceptualization, Investigation, Methodology, Project administration, Supervision, Writing – original draft, Writing – review & editing

