## Supplementary Materials for "Ketamine-Assisted Psychotherapy for Generalized Anxiety Disorder: A Comprehensive Case Report with Integrated Neurophysiological Imaging Using Magnetoencephalography"

### Cognitive-behavioural task descriptions

*Response inhibition* - The “Double Trouble” task is a test of focused attention and response inhibition. The right prefrontal cortex, and in particular the dorsolateral region, is involved in tests that require sustained focused attention.

*Attention* - The “Feature match” task measures visual processing by requiring the participant to judge small visual differences between pairs of overlapping triangles. Brain region activity associated with performance of this task includes the mid-ventrolateral frontal cortex and right inferior frontal gyrus

*Working memory* - The “Token search” task briefly displays elements of a grid-based pattern in sequence; after the pattern disappears, the participant is required to repeat the pattern using touch input. The difficulty of the task is adaptively modified by increasing or decreasing the number of elements in the sequence. Brain region activity associated with performance of this task includes the frontal and temporal lobes, amygdalo-hippocampal region, premotor cortex and dorsolateral and

*Visuospatial processing* - The “Polygons” task requires quick visuospatial processing, object recognition, and reasoning to determine if the shapes are the same. Brain region activity associated with performance of this task includes the intraparietal sulcus and right dorsolateral prefrontal cortex.

*Verbal reasoning* - In the “Grammatical reasoning” task a statement appears at the top of the screen, and two objects underneath. The subject’s task is to reason about the relationships among the objects and determine if the statement is true or false. Responding quickly and accurately is required for high scores. Grammatical Reasoning primarily involves two cognitive processes: verbal based reasoning to determine what the sentence should be describing, and comparison of this internally-generated answer with the image on the screen. Brain region activity associated with performance of this task includes the posterior temporal lobe, superior parietal lobe, dorsal and ventral prefrontal cortex.

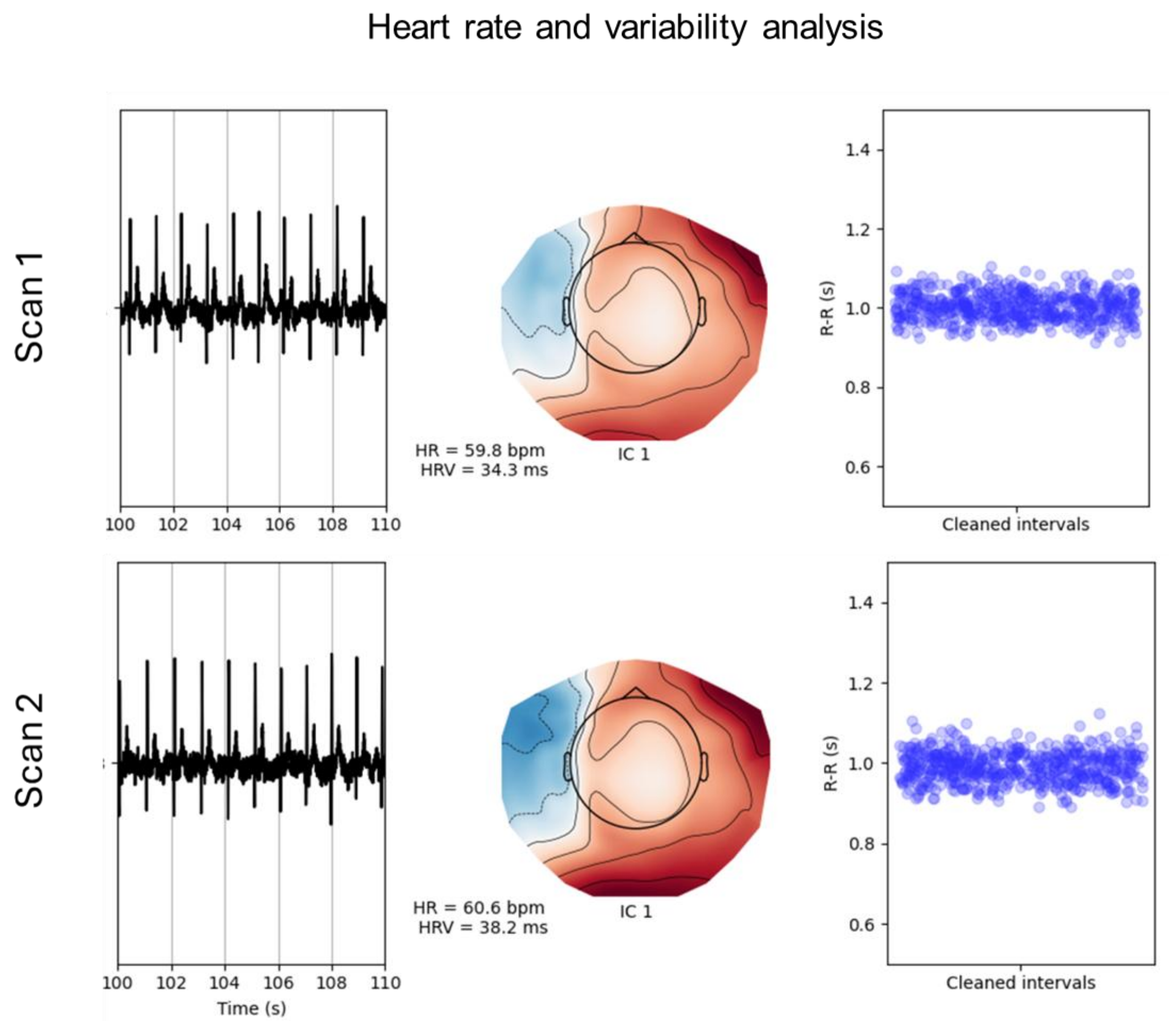


**Figure S1.** MEG scan resting heart rate and heart rate variability analysis. Representative data from two conditions showing cardiac and MEG-derived independent component (IC) activity. **Left:** Time series of cardiac activity over a 110-second interval. **Middle:** Scalp topography of an independent component (IC 1) associated with cardiac activity, with corresponding heart rate (HR) and heart rate variability (HRV) values. **Right:** Scatter plot of cleaned RR intervals (R-R_n in seconds) illustrating heart rate variability. The top and bottom rows represent different experimental conditions or subjects, with subtle differences in HR and HRV.

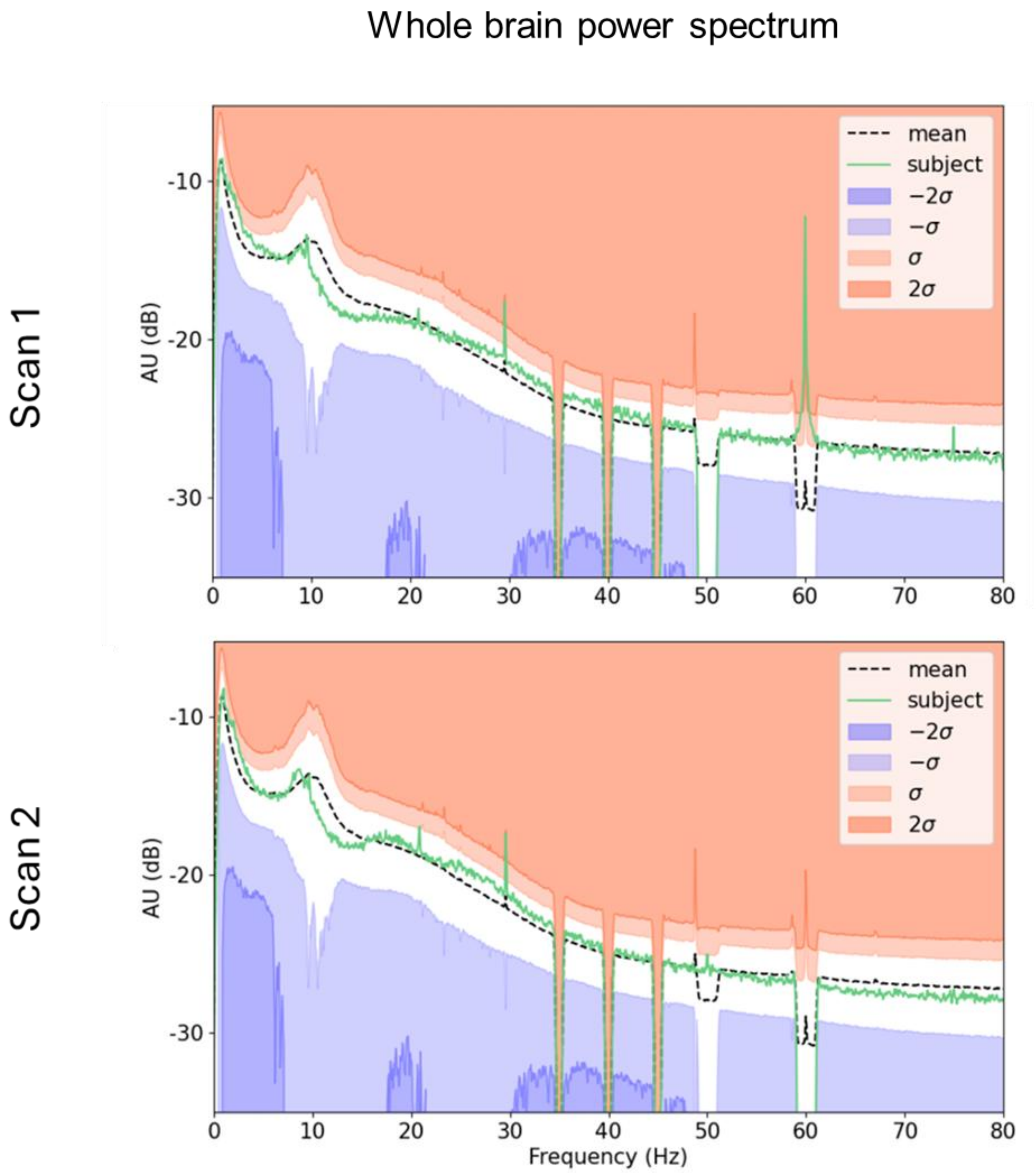


**Figure S2**. MEG scan mean whole-brain power spectrum. Power spectral density (PSD) plots comparing two conditions. The x-axis represents frequency (Hz), while the y-axis shows amplitude in arbitrary units (AU) on a decibel (dB) scale. The dashed black line represents the mean PSD across subjects, while the green line corresponds to an individual subject. Shaded regions indicate standard deviations (σ) from the mean, with blue representing negative deviations (−σ, −2σ) and red indicating positive deviations (σ, 2σ). Prominent peaks in the spectrum suggest residual artifacts or neural oscillatory activity at specific frequencies.
